# Has a Natural Endemic Focus for Dengue Been Established in Fujian Province,China? An Assessment Based on Four Core Evidence Dimensions, 2014–2024

**DOI:** 10.64898/2026.02.26.26347233

**Authors:** Shenggen Wu, Jinzhang Wang, Wenjing Ye, Yuedong Lin, Zhinan Guo, Yuwei Weng, Jun Han

## Abstract

**Background:** Dengue fever is a major neglected tropical disease with a rapidly rising global burden, and localized outbreaks are increasingly reported in southern subtropical China. Fujian Province, a coastal subtropical region with favorable ecological conditions for Aedes albopictus breeding and frequent cross-border exchanges with dengue-endemic areas, has had continuous local dengue cases for over a decade, raising concerns about the establishment of a stable natural endemic focus. Sustained local dengue transmission is defined by four core criteria, but no systematic assessment of these criteria has been conducted for Fujian using long-term multi-dimensional surveillance data. We aimed to evaluate whether a natural endemic focus for sustained local dengue transmission has been established in Fujian Province from 2014 to 2024 using four core evidence dimensions.

**Methods:** We extracted data on imported and locally acquired dengue cases in Fujian from 2014 to 2024 from China’s National Notifiable Disease Reporting System (NNDRS). Serological surveillance for dengue IgG antibodies and virological surveillance for dengue virus in Aedes albopictus were conducted at seven sentinel sites. The study period was stratified into three phases based on the impact of COVID-19 non-pharmacological interventions: pre-pandemic (2014–2019), pandemic(2020–2022), and post-pandemic(2023–2024). Descriptive epidemiological analysis and data visualization were performed using R software (version 4.4.1), with t-tests for continuous variables and χ² tests for categorical variables.

**Results:** A total of 3,606 dengue cases were reported in Fujian during the study period, including 1,229 imported and 2,377 locally acquired cases. Key findings were as follows: (1) Temporal distribution: Local dengue transmission was completely interrupted during the 2020–2022 COVID-19 pandemic (0 local cases, only 26 imported cases), and resumed at a low level in 2023–2024 (160 local cases). (2) Serology: The overall population dengue IgG antibody positivity rate was 4.2% (66/15,736), with no statistically significant difference between pre-epidemic (3.8%, 30/7,835) and post-epidemic seasons (4.5%, 36/7,901; P=0.48), and no year with a positivity rate exceeding 10%. (3) Vector surveillance: Only one dengue virus-positive sample was detected among 385,000 Aedes albopictus mosquitoes collected during routine surveillance (Taijiang District, Fuzhou, October 2017), with no viral nucleic acid detected in all other samples. (4) Age distribution: The mean age of locally acquired cases (46.1±19.8 years) was significantly higher than that of imported cases (35.8±11.2 years, P<0.001), and local cases were concentrated in the middle-aged group (40–60 years) with no child-dominant pattern observed.

**Conclusions:** Fujian Province has not established a stable natural endemic focus for sustained local dengue transmission, and imported cases are the primary driver of local outbreaks in the region. Strengthened surveillance and early management of imported cases, integrated vector control targeting Aedes albopictus, and targeted public health education are critical and essential strategies to prevent the establishment of a dengue natural endemic focus in Fujian and other subtropical coastal regions with similar epidemiological characteristics.

**Author Summary:** Dengue fever is a rapidly spreading neglected tropical disease worldwide, and southern China faces persistent threats of local transmission due to favorable ecological conditions for mosquito breeding and frequent cross-border travel. Fujian Province, a subtropical coastal region in southeastern China, has reported annual local dengue cases for over a decade, raising public health concerns about the potential establishment of a stable natural endemic focus—where the virus circulates sustainably without relying on imported cases.

To address this critical question, we conducted a comprehensive 11-year assessment (2014–2024) of dengue transmission in Fujian using four key evidence dimensions defined for identifying dengue endemic foci: the continuity of local cases independent of imported sources, population antibody levels, dengue virus detection in local mosquitoes (Aedes albopictus), and the age distribution of infected patients. We also leveraged the COVID-19 pandemic(2020–2022) as a unique natural experiment, during which strict travel restrictions drastically reduced imported dengue cases, to test whether local transmission could persist on its own.

Our findings showed that local dengue transmission in Fujian completely stopped during the COVID-19 pandemic and only resumed when cross-border travel and imported cases recovered, confirming local transmission is entirely dependent on imported virus sources. Additionally, the local population had a very low dengue antibody positivity rate (4.2%), dengue virus was detected in only one mosquito sample over 11 years of surveillance, and local cases were concentrated in middle-aged adults (not children—the typical group affected in endemic areas). Together, these results confirm that Fujian Province has not established a stable natural endemic focus for dengue fever.

While no endemic focus exists yet, Fujian remains at high risk of imported-driven local outbreaks due to its climate and cross-border exchanges. Our study highlights three critical strategies to prevent the future establishment of a dengue endemic focus in Fujian and other similar subtropical coastal regions: strengthening surveillance and early response for imported dengue cases, implementing targeted mosquito control measures during peak transmission seasons, and conducting public health education to raise awareness of dengue prevention. These evidence-based interventions are key to blocking the formation of sustained local dengue transmission and protecting regional population health.

## 1. Introduction

Dengue fever (DF), a mosquito-borne viral disease caused by the dengue virus (DENV) and transmitted primarily by Aedes albopictus and Aedes aegypti, remains one of the most pressing neglected tropical diseases (NTDs) of global public health concern, with no specific antiviral therapy and a vaccine with limited coverage[1–2]. The global burden of dengue has escalated dramatically over the past two decades: reported cases surged from 505,430 in 2000 to a record 14.6 million in 2024, with the disease now endemic in more than 100 countries and territories across tropical and subtropical regions[1]. A comprehensive analysis of global dengue burden from 1990 to 2021 revealed that incidence nearly doubled from 26.45 million to 58.96 million, with South Asia, Southeast Asia, and tropical Latin America bearing the heaviest burden—these regions also face rising disability-adjusted life-years (DALYs), particularly among children under 5 years old[2]. In China, dengue transmission has expanded northward in recent years, with localized outbreaks and sporadic local cases occurring annually during the summer and autumn months in southern subtropical provinces—including Guangdong, Yunnan, and Fujian—driven by favorable climatic conditions for Aedes mosquito breeding and increasing cross-border travel and trade[3]. Climate change is further exacerbating this trend: projected warming is expected to expand the geographic range of Aedes aegypti northward in China, prolong its seasonal activity, and increase the risk of dengue transmission, particularly in coastal regions like Fujian[4].

Fujian Province, a coastal subtropical region in southeastern China, is a key frontier for dengue prevention and control in the country. The province’s high temperatures, abundant rainfall, and dense human population create ideal ecological conditions for Aedes albopictus proliferation, and its frequent cross-border exchanges with dengue-endemic regions in Southeast Asia result in a continuous influx of imported dengue cases[5,6]. As demonstrated in Guangdong and Yunnan Province, temperature (especially daily mean temperature and daily temperature range) and imported cases are the primary drivers of local dengue outbreaks—these factors directly influence mosquito bite rate, transmission probability, and the virus’s external incubation period[7]. For more than a decade, local dengue cases have been reported annually in Fujian, raising critical concerns among public health authorities about the potential establishment of a stable natural endemic focus—a state of sustained local transmission independent of imported infection sources. Such an endemic focus would mark a pivotal shift in dengue epidemiology in the region, posing long-term challenges to disease control and increasing the risk of large-scale outbreaks.

Sustained local dengue transmission, the hallmark of an established natural endemic focus, can be assessed from four core epidemiological and virological dimensions[1,8–14]: (1) the persistent occurrence of local cases without direct links to imported sources; (2) a relatively high population positivity rate of dengue IgG antibodies, reflecting cumulative and widespread viral exposure; (3) the stable circulation of DENV in local Aedes vector populations; (4) a child-dominant age distribution of cases, driven by naive immunity in pediatric populations with no prior exposure to DENV. While these assessment dimensions are well-recognized, their systematic application to evaluate dengue endemicity in Chinese provincial regions—particularly using long-term, multi-dimensional surveillance data—remains limited. Prior studies on dengue in Fujian have focused on epidemiological characteristics and spatiotemporal clustering[9,10], but no research has yet conducted a comprehensive evaluation of whether the province meets the conditions for a natural endemic focus based on these four dimensions. This gap is notable given that dengue remains an imported disease in most parts of China, with local transmission entirely dependent on cross-border virus introduction[7], and climate-driven changes in vector ecology (e.g., prolonged breeding seasons) could alter this dynamic in the future[4,12]. In the Pearl River Delta—geographically and climatically similar to Fujian—future dengue risk is projected to expand significantly under climate change, underscoring the urgency of assessing endemic potential in subtropical coastal regions[12].

The COVID-19 pandemic (2020–2022) provided a unique natural experiment to test the sustainability of local dengue transmission in regions with sporadic local cases. Strict non-pharmacological interventions implemented during this period—including international travel restrictions, mandatory entry quarantine, and enhanced community surveillance—drastically reduced the number of imported dengue cases across China[11,13]. Similar observations were reported in Southeast Asian countries, where COVID-19 lockdowns altered human-mosquito contact patterns and temporarily disrupted local dengue transmission, though some regions (e.g., Singapore) later experienced resurgences due to relaxed restrictions and altered environmental conditions[6]. This unprecedented disruption of imported infection sources offers a critical opportunity to verify whether local transmission in Fujian could persist independently, a key criterion for an endemic focus.

Against this backdrop, we conducted a systematic assessment of dengue endemicity in Fujian Province using 11 years of comprehensive surveillance data (2014–2024), aligned with the four core criteria for sustained local transmission. We stratified the study period into three phases (pre-pandemic, pandemic, post-pandemic) to leverage the COVID-19 natural experiment, and integrated data from national notifiable disease reporting, population serological surveillance, Aedes vector virological testing, and case age distribution analysis. Our approach builds on previous work demonstrating the value of combining long-term surveillance data with climate and importation dynamics to understand dengue transmission in subtropical regions[4,7]. This study aims to provide evidence-based conclusions on the presence (or absence) of a dengue natural endemic focus in Fujian, and to propose targeted public health interventions to prevent the establishment of sustained local transmission. The findings not only fill a critical research gap in dengue surveillance in southeastern coastal China but also provide a replicable methodological framework for assessing dengue endemicity in other subtropical regions facing similar threats of imported virus introduction and vector-borne transmission.

## 2. METHODS

### 2.1 Data Collection

#### 2.1.1 Case Data

Data on imported and locally acquired dengue cases among residents of Fujian Province from 2014 to 2024 were extracted from China’s National Notifiable Disease Reporting System (NNDRS), a nationwide mandatory surveillance platform with standardized case reporting and verification protocols for infectious diseases in China. A case was defined as imported if the patient had a travel history to a dengue-endemic region within 14 days prior to symptom onset, with no evidence of potential infection in their county or district of residence. A case was classified as locally acquired if the patient had no travel history outside their resident county or district within 14 days before onset; for patients with such travel history, local acquisition was confirmed only after excluding the possibility of infection in dengue-endemic areas via epidemiological investigation.

#### 2.1.2 Serological Surveillance

Serological surveillance for dengue IgG antibodies was conducted at seven sentinel surveillance sites (3 national: Taijiang District, Lianjiang County, Shishi City; 4 provincial: Xiapu County, Jimei District, Hanjiang District, Jian’ou City) across Fujian Province (Figure 1), selected based on dengue transmission risk, population density, and cross-border travel frequency. At each sentinel site, 700 serum samples were collected annually from healthy community residents (excluding individuals with a history of dengue infection or vaccination) in two phases: May (pre-epidemic season) and November (post-epidemic season), corresponding to the pre-and post-peak periods of dengue transmission in southeastern China. Dengue IgG antibody detection was performed using a commercially available enzyme-linked immunosorbent assay (ELISA) kit, in strict accordance with the manufacturer’s standardized operating procedures (SOPs). All laboratory tests were conducted in certified microbiology laboratories of local Centers for Disease Control and Prevention (CDCs) to ensure quality control.

**Figure 1.**
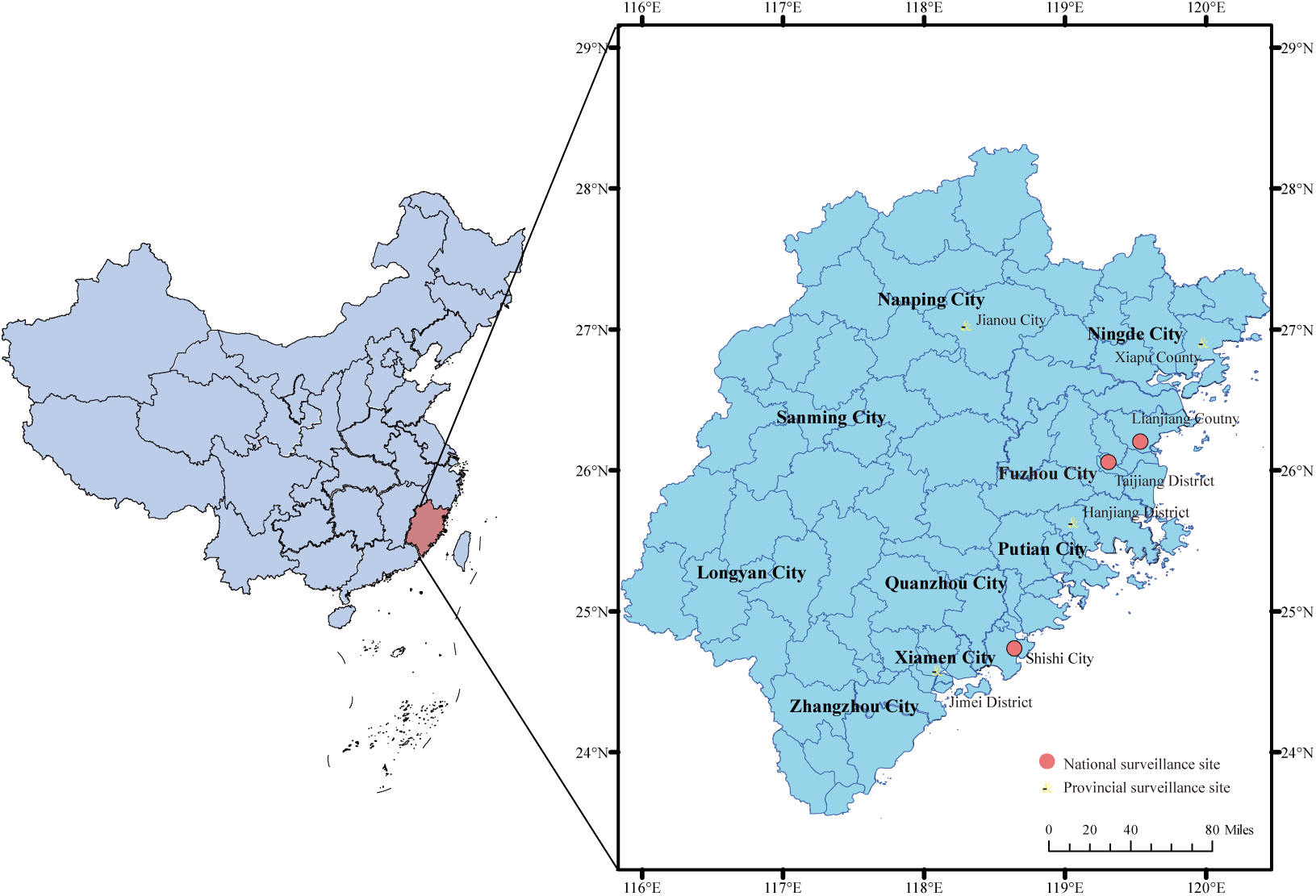
Geographic distribution of dengue serological and vector surveillance sites in Fujian Province, China Approval Number of Map Examination GS (2024) 0650

#### 2.1.3 Vector Surveillance

Entomological surveillance targeting Aedes albopictus—the primary dengue vector in Fujian Province—was conducted at the same seven sentinel sites during the annual peak dengue transmission season (June–October). At each site, a minimum of 100 adult Aedes albopictus mosquitoes were collected monthly using standard mosquito collection methods (human landing catches and ovitrap surveillance). Collected mosquitoes were homogenized in batches, and viral nucleic acid was extracted from homogenates using a viral RNA extraction kit. Nucleic acid amplification testing (RT-PCR) was used for the detection of dengue virus (DENV) nucleic acid, with universal primers targeting the conserved regions of the DENV genome to cover all four serotypes (DENV-1 to DENV-4). Negative and positive controls were included in each round of testing to validate assay accuracy and avoid false positive/negative results.

### 2.2 Study Design and Analysis

The 11-year study period (2014–2024) was stratified into three distinct phases based on the impact of COVID-19 non-pharmacological interventions (NPIs)—including international travel restrictions, mandatory entry quarantine, and enhanced community health surveillance—on cross-border travel and imported dengue transmission:1.Pre-pandemic phase(2014–2019): No COVID-19-related travel restrictions; normal cross-border travel and imported case influx.2.Pandemic phase (2020–2022): Strict nationwide COVID-19 NPIs with near-total suspension of non-essential international travel.3.Post-pandemic phase (2023–2024): Gradual lifting of COVID-19 travel restrictions; recovery of cross-border travel and resumption of imported case transmission.

### 2.3 Statistical analysis

Descriptive epidemiological analysis was performed to characterize the temporal distribution (annual and monthly) of imported and locally acquired dengue cases, population dengue IgG antibody positivity rates (overall, pre-vs. post-epidemic), DENV detection in Aedes albopictus, and age distribution of cases. For statistical comparisons, independent samples t-tests were used to analyze continuous variables (e.g., mean age of cases), and Pearson’s χ² tests were applied to compare categorical variables (e.g., dengue IgG antibody positivity rates between pre-and post-epidemic seasons).The map was generated using ArcGIS 10.2. All other data visualization (including case distribution curves, seropositivity trend plots, and age-structured pyramids) was generated using R software (version 4.4.1) with the ggplot2 package. A two-tailed P-value<0.05 was considered statistically significant for all analyses.

## 3. RESULTS

### 3.1 Temporal Distribution: Complete Interruption of Local Dengue Transmission During the COVID-19 Pandemic

A total of 3,606 dengue cases were reported in Fujian Province over the 11-year study period (2014–2024), comprising 1,229 imported cases and 2,377 locally acquired cases (Figure 2). Stratified analysis by the three COVID-19-associated phases revealed distinct transmission dynamics:

- Pre-pandemic phase (2014–2019): A steady influx of imported cases was recorded, with an annual average of 167 cases, accompanied by frequent local transmission that yielded an annual average of 377 locally acquired cases (range: 13–961 cases per year).
- Pandemic phase (2020–2022): Strict non-pharmacological interventions led to a drastic reduction in imported cases (annual average: 9 cases, total 26), and no locally acquired cases were reported during this period, marking a complete cessation of local dengue transmission.
- Post-pandemic phase (2023–2024): With the resumption of cross-border travel, imported cases rebounded to an annual average of 102 cases, and local transmission resumed at a low level, with an annual average of 80 locally acquired cases (range: 16–144 cases per year, total 160).

**Figure 2.**
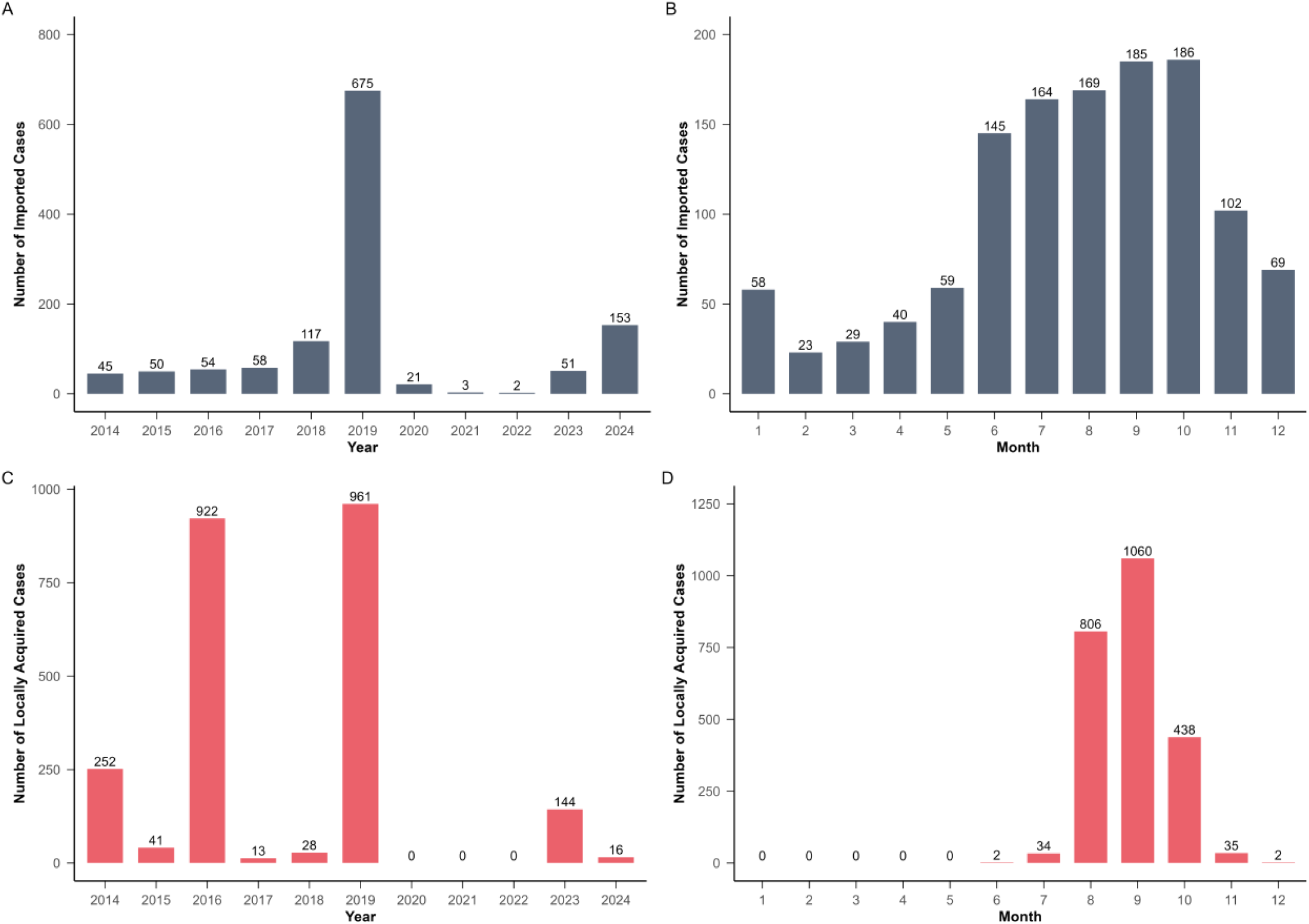
Temporal distribution of imported and locally acquired dengue cases in Fujian Province, 2014–2024. (A) Annual imported cases; (B) Monthly imported cases; (C) Annual locally acquired cases; (D) Monthly locally acquired cases

Monthly distribution analysis further characterized transmission patterns (Figure 2D): imported cases were detected year-round, with a clear peak in June–October (69.1%, 849/1,229). In contrast, locally acquired cases were restricted to the months of June–December, and the vast majority (96.9%, 2,304/2,377) occurred during the peak transmission window of August–October, consistent with the seasonal breeding activity of Aedes albopictus in southeastern China.

### 3.2 Serological Surveillance: Low and Non-Significant Population Dengue IgG Antibody Positivity

A total of 15,736 serum samples from healthy residents across the seven sentinel surveillance sites were tested for dengue IgG antibodies, yielding an overall provincial positivity rate of 4.2% (66/15,736) (Figure 3). Stratified analysis by transmission season showed a slightly higher positivity rate in the post-epidemic season (4.5%, 36/7,901) compared with the pre-epidemic season (3.8%, 30/7,835), but this difference was not statistically significant (χ² =0.498, P=0.48). Temporal trend analysis across the study period confirmed that the dengue IgG antibody positivity rate remained consistently low, with no single year recording a positivity rate exceeding 10% (Figure 3B).

**Figure 3.**
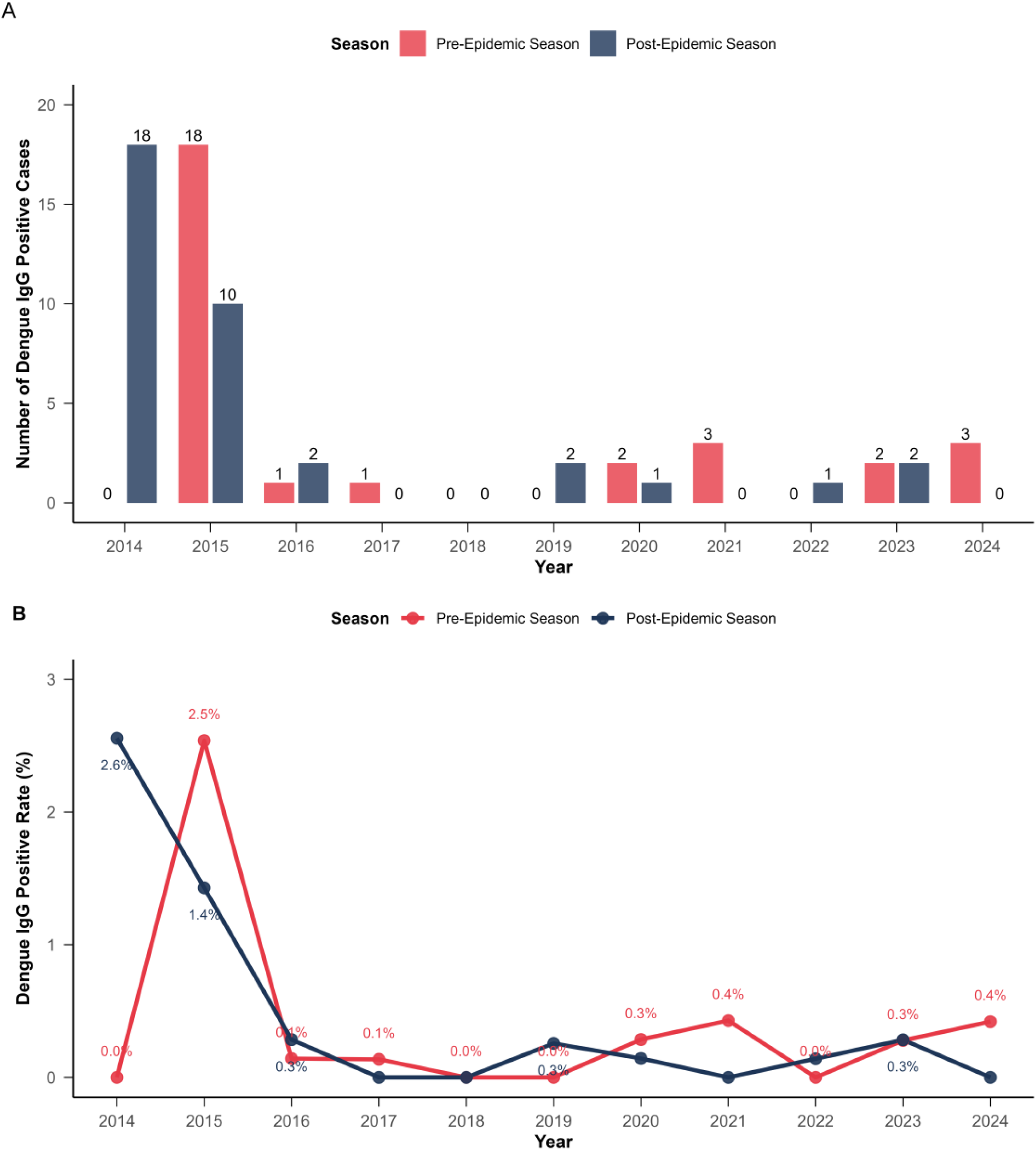
Serological surveillance of dengue IgG antibodies in healthy populations, 2014–2024. (A) Annual number of positive cases (pre-vs. post-epidemic); (B) Temporal trend of positivity rates

### 3.3 Vector Surveillance: Rare Dengue Virus Detection in Aedes albopictus With No Sustained Viral Circulation

Routine entomological surveillance was conducted at the seven sentinel sites during the annual peak dengue transmission seasons (2014–2024), with a total of 385,000 adult Aedes albopictus mosquitoes collected and tested for dengue virus nucleic acid. Viral RNA was detected in only one sample, which was obtained from the Taijiang District sentinel site in Fuzhou City in October 2017 and linked to the local dengue outbreak reported that year. No dengue virus nucleic acid was detected in any of the remaining mosquito samples across all surveillance years and sites, confirming the absence of stable, sustained dengue virus circulation in local vector populations.

### 3.4 Age Characteristics: Non-Child-Dominant Distribution of Locally Acquired Cases

The mean age of patients with locally acquired dengue cases (46.1 ± 19.8 years) was significantly higher than that of patients with imported cases (35.8± 11.2 years) (t=-19.41, P<0.001) (Figure 4A). This age disparity was consistent across the pre-pandemic and post-pandemic phases:

**Figure 4.**
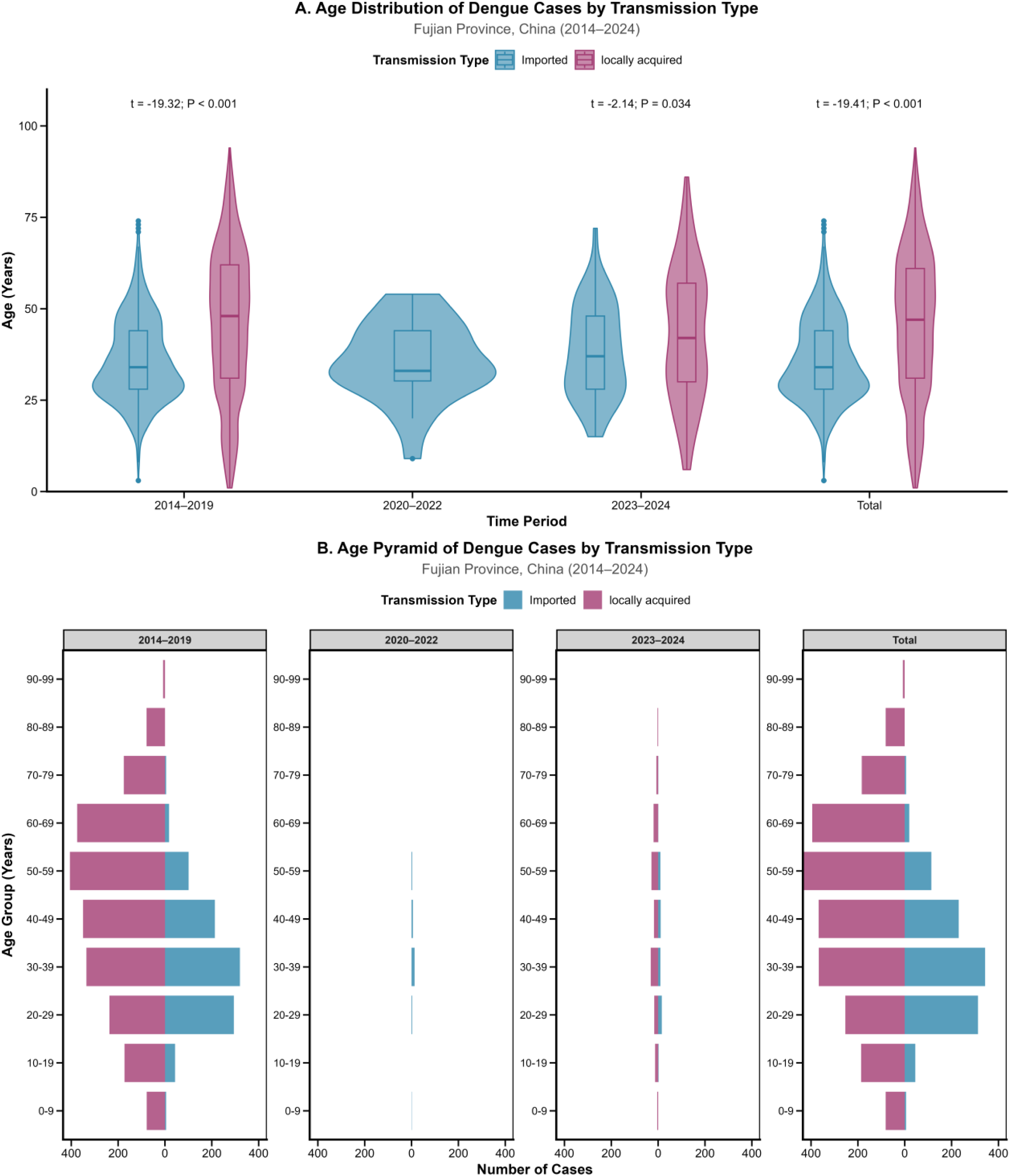
Age distribution of dengue cases by transmission type. (A) Mean age across time periods; (B) Age-structured pyramid (10-year groups)

2014–2019: Mean age of locally acquired cases (46.3 ± 19.9 years) vs. imported cases (35.7±11.2 years), (P<0.001).

2023–2024: Mean age of locally acquired cases (43.2 ± 18.0 years) vs. imported cases (38.2±12.7 years), (P=0.034).

Age-stratified analysis (10-year age groups) further demonstrated that locally acquired cases were predominantly concentrated in the middle-aged population (40–60 years). No predominance of cases was observed in pediatric or adolescent age groups (<18 years), and the age distribution pyramid confirmed a distinct absence of the child-dominant pattern characteristic of dengue-endemic regions (Figure 4B).

## 4. Discussion

This study provides a comprehensive, evidence-based evaluation of dengue endemicity in Fujian Province, southeastern China, using four core criteria for sustained local dengue transmission—temporal continuity of local cases independent of imported sources, population serological profile, vector-borne viral circulation, and case age distribution—with 11 years of systematic surveillance data(2014–2024). The findings converge on a definitive conclusion: **Fujian Province has not established a stable natural endemic focus for dengue fever, and all local transmission events in the region are driven by imported infection sources.** This conclusion is robustly supported by consistent evidence across all four assessment dimensions, and further validated by the unique natural experiment of COVID-19 non-pharmacological interventions (NPIs), which disrupted imported dengue transmission and led to a complete cessation of local cases in the province.

First, the interruption of local dengue transmission during the 2020–2022 COVID-19 pandemic represents the most compelling evidence for the absence of sustained local transmission in Fujian. Strict nationwide travel restrictions, mandatory entry quarantine, and enhanced border health surveillance reduced annual imported dengue cases from a pre-pandemic average of 167 to just 9, eliminating the primary source of viral introduction into the province. The subsequent complete disappearance of locally acquired cases during this period, and their resurgence in 2023–2024 coinciding with the recovery of cross-border travel and imported case numbers, directly demonstrates that local transmission in Fujian is entirely dependent on imported DENV. This finding aligns with similar observations from other southern Chinese provinces (e.g., Guangdong)[11], where COVID-19 NPIs also halted local dengue transmission, confirming that sustained, imported-independent local circulation has not yet been achieved in mainland China’s subtropical regions. Notably, the low volume of local cases in the post-pandemic phase (annual average 80) further indicates that even with resumed imported transmission, viral spread in Fujian remains limited and non-sustained, lacking the community transmission potential required for endemic focus formation.

Second, the low overall population dengue IgG antibody positivity rate of 4.2% (66/15,736) in Fujian strongly corroborates the non-endemic status of the province, and stands in stark contrast to the high seroprevalence observed in established dengue-endemic regions worldwide. WHO guidelines and global epidemiological studies confirm that repeated viral exposure in endemic areas results in dengue IgG seropositivity rates typically exceeding 50%, and often reaching >70% in highly endemic settings[12,14]. For example, a population-based study in the Philippines—a major dengue-endemic country—reported an IgG seropositivity rate of 76.2% (95% CI: 71.9–80.0) [15], more than 18 times the rate observed in Fujian. Globally, the burden of dengue is highest in middle and low-middle Socio-Demographic Index (SDI) regions, where seroprevalence rates reflect cumulative exposure to repeated outbreaks [2]. In these regions, seropositivity often exceeds 50% by adolescence[14], a pattern not observed in Fujian. Furthermore, the lack of a statistically significant difference in seropositivity between pre-epidemic (3.8%) and post-epidemic (4.5%) seasons in Fujian (P=0.48), and the absence of any year with a positivity rate exceeding 10%, reflect minimal cumulative population exposure to DENV. This low seroprevalence rules out widespread, undetected community transmission in the province, and confirms that the local population remains largely naive to dengue virus—an important characteristic of non-endemic regions, and a critical distinction from endemic areas where herd immunity shapes transmission dynamics.

Third, the near-absence of dengue virus detection in local Aedes albopictus populations eliminates a key prerequisite for the establishment of a natural endemic focus: stable, sustained viral circulation in the primary vector. Over 11 years of routine entomological surveillance, only a single DENV-positive Aedes albopictus sample was identified (Taijiang District, Fuzhou, October 2017), and subsequent virological investigations confirmed this positive sample was directly associated with the local dengue outbreak reported in Fujian that year[16]. No viral nucleic acid was detected in the remaining 384,999 tested mosquitoes across all surveillance sites and years, confirming that DENV does not persist in Fujian’s vector populations between imported transmission events. This is consistent with studies highlighting the importance of meteorological factors (e.g., temperature, rainfall) in vector viral circulation: rainfall (particularly summer peak rainfall) was found to be more critical than temperature in driving mosquito population growth and viral transmission[5]. A five-stage mathematical model of Aedes albopictus population dynamics demonstrated that seasonal rainfall peaks—rather than temperature distribution—are the strongest predictor of mosquito abundance, which directly influences viral circulation potential[5]. The lack of such persistent vector-borne viral circulation in Fujian means the province’s vector population cannot act as a viral reservoir, precluding the formation of a self-sustaining endemic focus. Moreover, future climate change may alter this dynamic: projected warming is expected to increase the number of annual life-cycle completions of Aedes aegypti in coastal China, potentially enhancing vector-borne viral circulation and elevating the risk of endemic focus formation [4].

Fourth, the age distribution of locally acquired dengue cases in Fujian—dominated by middle-aged adults (40–60 years) with a mean age of 46.1±19.8 years, and no pediatric/adolescent predominance—aligns with the classic age pattern of non-endemic or emerging dengue regions, and directly contrasts with the child-dominant distribution characteristic of established endemic areas[12]. In endemic regions such as Southeast Asia and Latin America, children and adolescents account for the majority of dengue cases due to cumulative viral exposure in older age groups, leaving pediatric populations with naive immunity and high susceptibility [12]. Globally, children under 5 years of age bear the highest DALYs burden from dengue, though incidence is often highest in adolescents and the elderly[2]. This age pattern—children as the most vulnerable to severe outcomes—further distinguishes endemic regions from Fujian, where local cases are concentrated in adults and severe outcomes are rare. In contrast, the adult-dominant case distribution in Fujian reflects transmission dynamics typical of imported-driven dengue: viral spread is limited to adult populations with higher exposure to imported cases (e.g., through cross-border travel, workplace contact, or community interactions with returning travelers), and does not reach pediatric groups—an indication that transmission is not sustained enough to permeate the broader community. This age pattern was consistent across both the pre-pandemic and post-pandemic phases, further validating that imported-driven transmission, rather than endemic circulation, shapes dengue epidemiology in Fujian.

### Limitations

This study has three notable limitations that should be considered when interpreting its findings. First, serological and vector surveillance were conducted exclusively at seven high-risk sentinel sites across Fujian Province, selected based on dengue transmission history, population density, and cross-border travel frequency. This targeted sampling may overestimate the provincial average dengue IgG antibody positivity rate, as surveillance sites were chosen for their higher dengue risk, and results may not be fully representative of low-risk areas of the province where exposure to DENV is likely even lower. Second, dengue antibody testing was performed using a commercial ELISA kit for IgG detection only, with no further serotyping or neutralizing antibody testing conducted. This limits our ability to characterize the specific DENV serotypes circulating in Fujian, and to distinguish between primary and secondary infections—information that would add nuance to our understanding of local transmission intensity and population immunity. Given that infection with different DENV serotypes can influence disease severity and immunity dynamics[2], and that serotype shifts are associated with outbreaks in endemic regions[2], future studies should incorporate serotyping to better characterize local transmission risks. Third, the study relied on reported dengue cases from China’s National Notifiable Disease Reporting System (NNDRS), which does not capture asymptomatic. Asymptomatic infections are known to account for a large proportion of dengue cases globally (up to 75% in some settings)[17], and their exclusion from our analysis may underestimate the true transmission intensity in Fujian, and potentially the level of population exposure to DENV. Additionally, we did not explicitly account for socioeconomic factors (e.g., urbanization, population density) or climate change projections, which have been shown to influence dengue transmission risk in similar coastal regions[4,8,12]. The Pearl River Delta’s projected expansion of dengue risk under climate change[8] highlights the need to integrate such factors in future assessments of Fujian’s endemic potential.

### Public Health Implications

While Fujian has not established a dengue natural endemic focus, the province remains at high and persistent risk of imported-driven local outbreaks, due to a confluence of ecological, demographic, and geopolitical factors. Fujian’s subtropical coastal climate—characterized by high temperatures and abundant rainfall year-round—creates ideal breeding conditions for Aedes albopictus, the primary dengue vector in the province[9,10]. As demonstrated in neighboring regions, temperature (particularly daily mean temperature and daily temperature range) directly influences mosquito vector parameters (e.g., bite rate, transmission probability) and thus dengue transmission risk[7]. A study of the 2019 dengue outbreaks in Guangzhou and Jinghong found that temperature-driven changes in mosquito behavior (e.g., increased biting rate) were the primary drivers of outbreak intensity, alongside imported cases[7]. Additionally, the province’s frequent cross-border travel and trade with dengue-endemic regions in Southeast Asia ensure a continuous influx of imported DENV cases, and the long incubation period of dengue virus (2–14 days)[18] and high prevalence of asymptomatic infections[17] make early detection and isolation of imported cases challenging. These factors create a constant risk of local transmission amplification, and highlight the need for sustained, targeted public health interventions to prevent the eventual establishment of a dengue endemic focus in Fujian. Furthermore, future climate change may exacerbate this risk by expanding vector distribution, prolonging transmission seasons, and increasing viral replication rates in mosquitoes[4,12], underscoring the need for proactive adaptation strategies.

To mitigate this risk, three core, evidence-based interventions are recommended for dengue prevention and control in Fujian Province, and are broadly applicable to other subtropical coastal regions of China and the globe facing similar imported dengue threats:

1. Strengthen imported case surveillance and early response: Enhance port health quarantine and post-arrival health monitoring for travelers returning from dengue-endemic regions, with a focus on real-time reporting and rapid epidemiological investigation of suspected cases. This aligns with findings from Guangzhou and Jinghong, where reducing the importation coefficient (via targeted quarantine and surveillance) significantly lowered local dengue cases [7]. This will reduce the window of viral transmission from imported cases to local vectors and populations.
2. Implement integrated, seasonally targeted vector control: Prioritize Aedes albopictus control measures during the annual peak dengue transmission season (June–October) in Fujian, including source reduction (elimination of artificial breeding sites), larvicide application, and adult mosquito control in high-risk areas (e.g., urban centers, border regions, and surveillance sites).Yang et al. further verified in Guangzhou that targeted control measures against Aedes albopictus during meteorologically driven population peaks could significantly reduce vector density and interrupt local dengue transmission chains[19]. Routine vector density monitoring should be maintained to identify and respond to potential transmission hotspots, with a focus on meteorological triggers (e.g., heavy rainfall) that drive vector abundance[5].
3. Conduct targeted public health education and community engagement: Develop and disseminate dengue prevention education tailored to high-risk populations (e.g., frequent international travelers, healthcare workers, and residents of urban coastal communities), focusing on recognizing dengue symptoms, reducing mosquito exposure, and reporting suspected cases promptly. Community participation in vector control (e.g., household breeding site elimination) is critical to achieving sustainable reduction in Aedes populations. Such community engagement is particularly important in the post-COVID-19 era, where changes in human mobility and contact patterns (e.g., increased indoor activity) may alter dengue transmission dynamics[20–22]. Additionally, education should emphasize the role of imported cases in driving local outbreaks, as awareness of travel-related risks can improve early reporting and reduce transmission.

## 5. Conclusions

This study provides the first systematic and evidence-based assessment of whether a natural endemic focus for sustained local dengue transmission has been established in Fujian Province, a subtropical coastal region in southeastern China, using 11 years of comprehensive surveillance data (2014–2024) and four internationally recognized core evaluation dimensions. Convergent evidence from the COVID-19-induced interruption of local transmission, a persistently low population dengue IgG antibody positivity rate, the near-absence of sustained dengue virus circulation in local Aedes albopictus vectors, and a non-child-dominant age distribution of locally acquired cases confirms a definitive conclusion: Fujian Province has not established a stable natural endemic focus for dengue fever. All local dengue transmission events in the province are driven by imported infection sources, and the resumption of local cases in the post-pandemic period is directly associated with the recovery of cross-border travel and the rebound of imported dengue cases.

Fujian’s status as a non-endemic region for sustained dengue transmission highlights the critical role of targeted and sustained public health interventions to prevent the establishment of a natural endemic focus in the future. The province’s subtropical climate creates permanent ecological conditions favorable for Aedes albopictus breeding, and frequent cross-border exchanges with dengue-endemic regions in Southeast Asia ensure a continuous influx of imported viral sources—two factors that place Fujian at a persistent high risk of imported-driven local outbreaks. Without effective control measures, the cumulative introduction of dengue virus could eventually lead to sustained local circulation and the formation of an endemic focus, posing long-term challenges to regional public health security.

Against this backdrop, three core evidence-based strategies are indispensable for dengue prevention and control in Fujian Province: strengthened surveillance and early management of imported cases to block the primary viral introduction route; integrated and seasonally targeted vector control measures centered on Aedes albopictus to eliminate transmission vectors; and targeted public health education to improve community awareness and participation in dengue prevention. These interventions are not only applicable to Fujian but also provide a replicable reference for other subtropical coastal regions in China and across the globe that face similar threats of imported dengue transmission and have ecological conditions suitable for Aedes mosquito proliferation.

Long-term continuous surveillance is also essential to monitor the dynamic changes in dengue transmission in Fujian Province. Future research should expand surveillance coverage beyond the current seven sentinel sites to include rural and low-risk areas, combine serotyping and neutralizing antibody testing to characterize dengue virus circulation in the region, and incorporate asymptomatic infection surveillance to more accurately assess the true transmission intensity. Such efforts will provide a more comprehensive evidence base for optimizing dengue prevention and control strategies and safeguarding population health in southeastern China.

### Ethics Approval and Consent to Participate

This study utilized de-identified routine surveillance data of dengue fever, including case data from China’s National Notifiable Disease Reporting System (NNDRS), population serological surveillance data, and vector surveillance data, which were collected and managed by Fujian Center for Disease Control and Prevention and relevant municipal CDCs in Fujian Province in accordance with national infectious disease surveillance regulations of the People’s Republic of China.

Since the study only used secondary de-identified public health surveillance data without any identifiable personal information or intervention on human subjects, no ethical approval and informed consent were required according to the Ethical Review Guidelines for Biomedical Research Involving Human Subjects (National Health Commission of China, 2016) and the relevant ethical review regulations of Fujian Center for Disease Control and Prevention. All data collection and analysis procedures were conducted in compliance with national and local public health surveillance norms.

## Data Availability

This study utilized de-identified routine surveillance data of dengue fever, including case data from China’s National Notifiable Disease Reporting System (NNDRS), population serological surveillance data, and vector surveillance data, which were collected and managed by Fujian Center for Disease Control and Prevention and relevant municipal CDCs in Fujian Province in accordance with national infectious disease surveillance regulations of the People’s Republic of China. Since the study only used secondary de-identified public health surveillance data without any identifiable personal information or intervention on human subjects, no ethical approval and informed consent were required according to the Ethical Review Guidelines for Biomedical Research Involving Human Subjects (National Health Commission of China, 2016) and the relevant ethical review regulations of Fujian Center for Disease Control and Prevention. All data collection and analysis procedures were conducted in compliance with national and local public health surveillance norms.

